# Optimizing *Shigella* isolation: A multi-site evaluation of laboratory culture methods for *Shigella* detection, speciation, and serotyping with different transport media and sample types in the Enterics for Global Health study

**DOI:** 10.1101/2025.09.02.25334652

**Authors:** Taufiqur Rahman Bhuiyan, Jie Liu, Jane Juma, Bri’Anna Horne, Aneeta Hotwani, Henry Badji, Victor Adrian Maiden, John Benjamin Ochieng, Lucero A. Romaina-Cachique, Laura Riziki Aluoch, Lilian Achieng Ambila, Fatima Aziz, Mary Charles, Erika L. Feutz, Paul F. Garcia-Bardales, M. Jahangir Hossain, Junaid Iqbal, Taufiqul Islam, Abdoulie Jabang, Sheikh Jarju, Khuzwayo C. Jere, Furqan Kabir, Flywell Kawonga, Adama Mamby Keita, Farhana Khanam, Katia Manzanares Villanueva, Parvej Mosharraf, Richard Omore, Maribel Paredes Olortegui, Patricia B. Pavlinac, James A. Platts-Mills, Elizabeth T. Rogawski McQuade, Queen Saidi, Khandra Sears, Milagritos D. Tapia, Awa Traore, Firdausi Qadri, Pablo Penataro Yori, Farah Naz Qamar, Samba O. Sow, Karen L. Kotloff, Sharon M. Tennant, Eric R. Houpt, Jennifer Cornick, Ousman Secka, the EFGH Consortium

## Abstract

*Shigella* is a leading cause of diarrhea and dysentery in children under 5 in low-resource settings and several vaccines are in development. Due to its fastidious nature, *Shigella* can be difficult to culture and eventual vaccine trials will need to optimize the isolation of *Shigella* to ensure efficient sample sizes. In the recently concluded Enterics for Global Health study (EFGH) *Shigella* Surveillance study, we compared *Shigella* culture isolates rates between rectal swab vs. whole stool; between two swabs vs. one, and between Cary Blair (CB) vs. modified-Buffered Glycerol Saline (mBGS) transport media to identify the optimal methods for *Shigella* recovery by microbiologic culture. Among 9,476 children aged 6–35 months enrolled in the EFGH study from seven country sites, *Shigella* isolation rates did not differ significantly between CB (7.8%) and mBGS (7.9%) (p=0.545). Using two swabs improved detection rates (9.3%) compared to one, (7.9%) (p<0.001). Among the 2048 children from Bangladesh and the Gambia where both rectal swabs and whole stool were collected from the same children, rectal swabs were found to be non-inferior to whole stool for *Shigella* culture (12.4% and 12.7%, respectively with a difference of -0.29% (95% confidence interval -0.83% to 0.24). To optimize *Shigella* recovery for future multi-country vaccine trials, we recommend collecting two flocked rectal swabs in CB or mBGS media with strict adherence to transit conditions—an approach proven feasible across EFGH sites.

## Introduction

*Shigella* species (spp.) are a leading cause of moderate-to-severe diarrhea among children under five years of age, contributing to more than 80,000 child deaths per year [1]. *Shigella* is divided into four spp. (*S. flexneri*, *S. sonnei*, *S. boydii* and *S*. *dysenteriae*) with the majority of shigellosis caused by *S. flexneri* and *S. sonnei* [2]. Although, *S. flexneri* exists as multiple serotypes, of which *S. flexneri* 2a, 3a, and 6 are the most common globally, there is geographic heterogeneity in burden and serotype/subserotype distribution [2-4]. Based on global burden data, it is estimated that a vaccine targeting *S. flexneri* 2a, 3a, and 6, and *S. sonnei,* could prevent more than 80% of shigellosis cases globally [2]. Multiple vaccine candidates are under development, and it is expected that at least one candidate will be tested in a Phase III licensure trial within the next five years [5].

Stool culture has long been considered the gold standard method for diagnosis of shigellosis and it remains a prerequisite step for downstream speciation, serotyping, and antimicrobial susceptibility testing [6]. However, the sensitivity of *Shigella* culture varies by disease severity and stage, with higher culture positivity in severe cases and optimal detection when stool is sampled early in illness [7, 8]. Other variables that may affect stool culture sensitivity include laboratory capacity, sample type, handling conditions, transportation time and the type of media used to preserve samples prior to plating [9].

Rectal swabs are used for *Shigella* culture when whole stool is unavailable or when a prompt sample collection is required prior to antibiotic administration. Several studies in Europe, the Americas and Asia [9-14] which have compared rectal swabs to whole stool for diagnostic performance of enteric pathogens causing diarrhoea have consistently shown that both methods are comparable with no significant differences [10, 12]. Although data from Africa are limited, studies conducted in Rwanda and Botswana [15-17] have shown that rectal swabs can serve as a practical alternative to bulk stool, particularly in settings where stool collection may be challenging and rapid diagnostics are essential. Use of flocked rectal swabs is an important development for enteropathogen identification when whole stool is not available [10]. Notably, rectal swabs have demonstrated superior performance for molecular testing [15]. However, no studies using culture methods have yet directly compared *Shigella* isolation from rectal swabs and whole stool in high-burden settings, therefore the current study was conducted to address this gap.

Transport media is used when fecal samples cannot be cultured immediately as the fastidious nature of *Shigella* makes it sensitive to suboptimal conditions. Cary Blair (CB) medium is the most widely used medium for fecal sampling due to its ability to preserve bacterial viability during transport [18]. Its use, however, has not been studied in low- and middle-income countries (LMICs), where variations in storage conditions, particularly in rural settings with limited infrastructure, can negatively affect sample quality and, consequently, diagnostic accuracy [19]. Buffered glycerol saline (BGS) is an alternative transport medium with some evidence of it yielding a higher *Shigella* culture positivity than CB, albeit in small studies [20, 21]. Additionally, one study showed that a modified version of BGS (0.5% agar and reduced glycerol to 15%) showed enhanced recovery of *Shigella* [22].

As vaccine trials are planned with the goal of preventing culture-confirmed *Shigella* infections, optimizing detection methods from fecal samples will be critical to the success of *Shigella* vaccine development. In the seven-country Enterics for Global Health (EFGH) *Shigella* surveillance study which enrolled children aged 6-35 months seeking care for diarrhea, we aimed to identify the optimal sample type and transport media for *Shigella* stool culture. Findings from this study will directly inform laboratory protocols for future *Shigella* licensure trials.

## Materials and Methods

The EFGH *Shigella* surveillance study protocol is described in detail elsewhere [23, 24]. In brief, diarrheal cases were enrolled from public health facilities and hospitals in seven countries: Malawi, Kenya, Mali, The Gambia, Bangladesh, Pakistan, and Peru. After verbal screening consent was obtained from caregivers, children between the ages of 6-35 months attending EFGH health facilities with diarrhea, defined as three or more loose or watery stools in the 24 hours prior to enrollment, were screened during EFGH working hours, using a standardized questionnaire. Written informed consent for the study was obtained from caregivers of eligible children after screening. Other eligibility criteria were listed previously [24]. Detailed clinical demographic information was collected through standardized case report forms and physical examinations.

Two nylon flocked rectal swabs (COPAN^©^, USA) were collected from enrolled children immediately after screening, or as close to screening as possible preferably prior to antibiotic administration (if prescribed). Whole stool was also collected from children in two of the seven EFGH country sites (Bangladesh and The Gambia) if produced within six hours of screening and prior to the child being ready to return home. One rectal swab was immediately placed in CB medium and the other in mBGS. The rectal swab order for each respective media was swapped halfway through the study to avoid concern that order of swabbing influenced the quantity of fecal sample available to culture. To ensure whole stool was prepared similarly to rectal swabs for the swab/stool comparison, a nylon flocked rectal swab was used to touch the stool targeting bloody, slimy, mucoid, or watery areas and placed into the appropriate tubes with CB transport media (first half of the study period) or mBGS media (second half of study period).

Immediately after collection, all fecal samples were placed in a cold box and maintained at 2-8°C monitored for temperature excursions using a single-use 2-8°C temperature monitors (WarmMark, Spotsee company or 3M). Samples were transferred to an in-country central laboratory within 16 hours of collection. Upon reception, an accessioning form was completed and samples eligible for processing were cultured for *Shigella* isolation. The microbiological culture process has been described in detail previously [24]. In brief, swabs in mBGS and CB were streaked on MacConkey (MAC) and Xylose Deoxycholate (XLD) agar and incubated for 18-24 hours at 35-37°C. Up to 10 suspected *Shigella* non-lactose fermenter (NLF) colonies were subcultured on trypticase soy agar (TSA) and screened using a series of biochemical tests. Pure NLF colonies from the TSA were subcultured on Triple sugar Iron (TSI), Motility indole orthenine (MIO) or Motility indole urea (MIU), urea, Lysine Decarboxylase, and Tryptic soy agar (TSA) and incubated as described previously [24]. A colony was assigned as suspected *Shigella* if it exhibited typical biochemical characteristics and subjected to latex agglutination. Pure, fresh suspected *Shigella* colonies on TSA were serotyped using polyvalent and monovalent antisera. Polyvalent antisera were used to identify *S. dysenteriae*, *S. flexneri*, *S boydii*, and *S. sonnei.* If a colony was identified as *S. flexneri*, monovalent antisera were used to identify the *S. flexneri* serotype as previously described [24].

To harmonize methods across different sites, a joint working group was convened comprising representatives from coordinating and implementing teams. The group developed a set of consensus standard operating procedures (SOPs), worksheets, and data collection forms for every component of the laboratory methodology, including sample collection and transport, media preparation, and steps for primary *Shigella* isolation, biochemical identification, serotyping with agglutination assays, and susceptibility testing [24]. Essential laboratory supplies were procured centrally as needed. The team followed quality control (QC) and quality assurance procedures along with site visits approximately every 6 months to review the implementation and adherence to protocols.

The protocol for this study was approved by the respective Institutional Review Boards (IRBs) for each country study site and their affiliates. This study was conducted according to Good Clinical Practice (GCP), including Good Clinical Laboratory Practice (GCLP), the Declaration of Helsinki, IRB and local rules and regulations specific to each EFGH country.

### Statistical Analysis

The percentage of rectal swab samples from each media that isolated *Shigella* were summarised by serotype and country site and two-sided 95% confidence intervals were calculated assuming a binomial distribution. McNemar’s test of superiority was employed to assess whether either medium had significantly different *Shigella* isolation proportions based on p<0.05.

The percentage of whole stools and rectal swabs that isolated *Shigella* were calculated both overall and at each country site and further stratified by *Shigella* species. Because two swabs were collected for each participant, we considered only the *Shigella* isolation from the first rectal swab collected from each child which matches the media type that same child’s whole stool was transported in. Separately, a sensitivity analysis was performed to compare *Shigella* isolation from either of two rectal swabs to whole stool. To determine whether rectal swabs were inferior to whole stool for isolating *Shigella,* McNemar’s test of non-inferiority was used. We computed a one-sided 95% confidence interval for the absolute difference in proportions between sample types. The lower bound of the confidence interval was compared to the non-inferiority margin of an absolute difference of 0.01, or 1%, to establish non-inferiority.

## Results

The demographic characteristics of the 9,476 children aged 6-35 months enrolled in the EFGH study are listed in **Table 1**. The majority of children fell in the 12-23 month age range (46.1%) or the 6-11 month age range (36.7%). The proportion of enrolled cases presenting with dysentery ranged from 6.6% in Mali to 17.2% in The Gambia. Dehydration and hospitalization also varied substantially between countries. Swabs were collected from all enrolled children in EFGH and whole stool from 1,097 (80.6%) children in Bangladesh and 975 (69.7%) children in The Gambia. Fourteen whole stool samples were excluded in analyses due to having been collected more than six hours from rectal swab collection. Fecal samples were placed in transport media on average 0-2 minutes after collection and average time between collection and arrival at the central laboratory for primary plating ranged from 1.4 hours in The Gambia to 4.4 hours in Malawi.

**Table 1:**
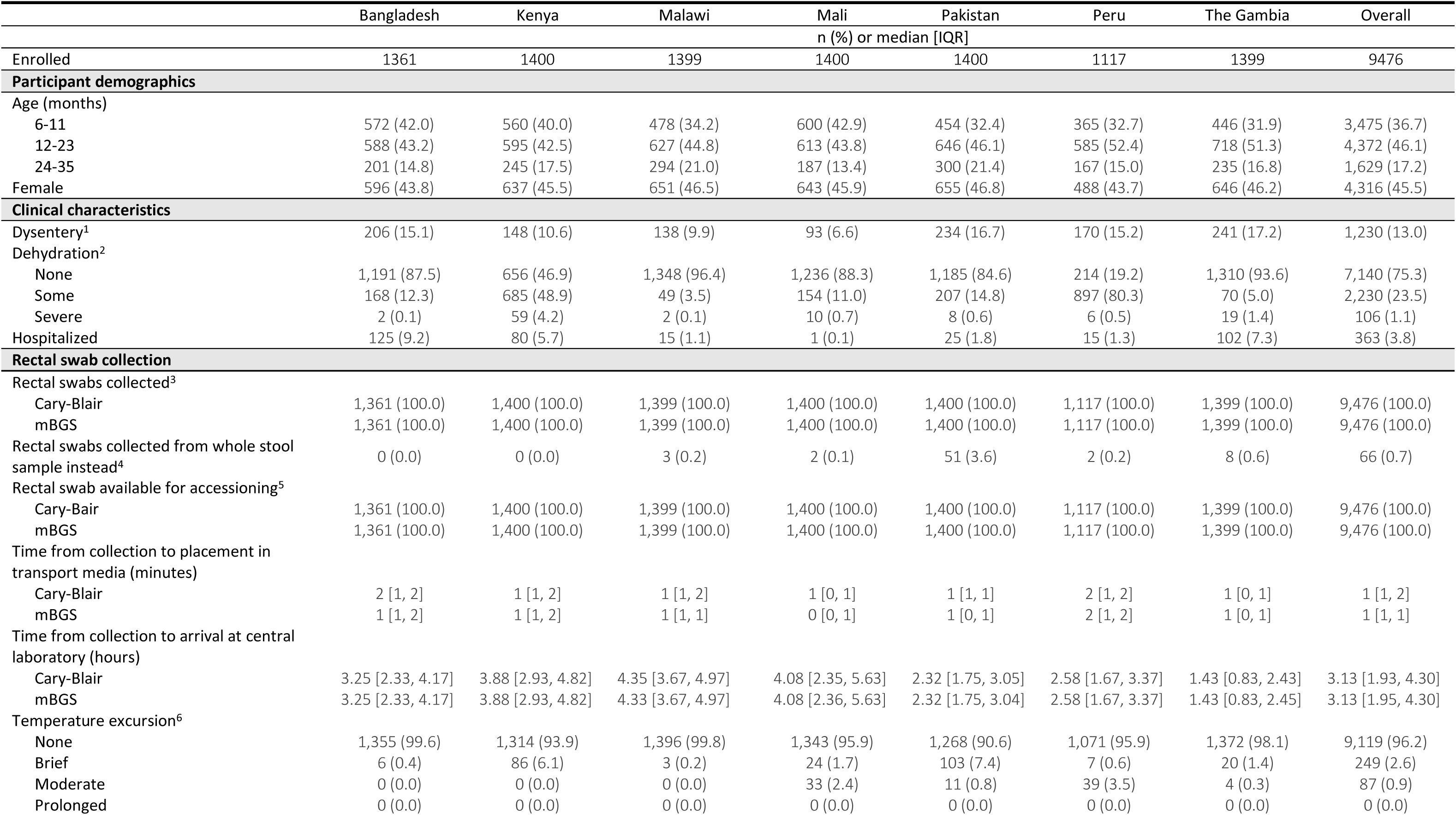
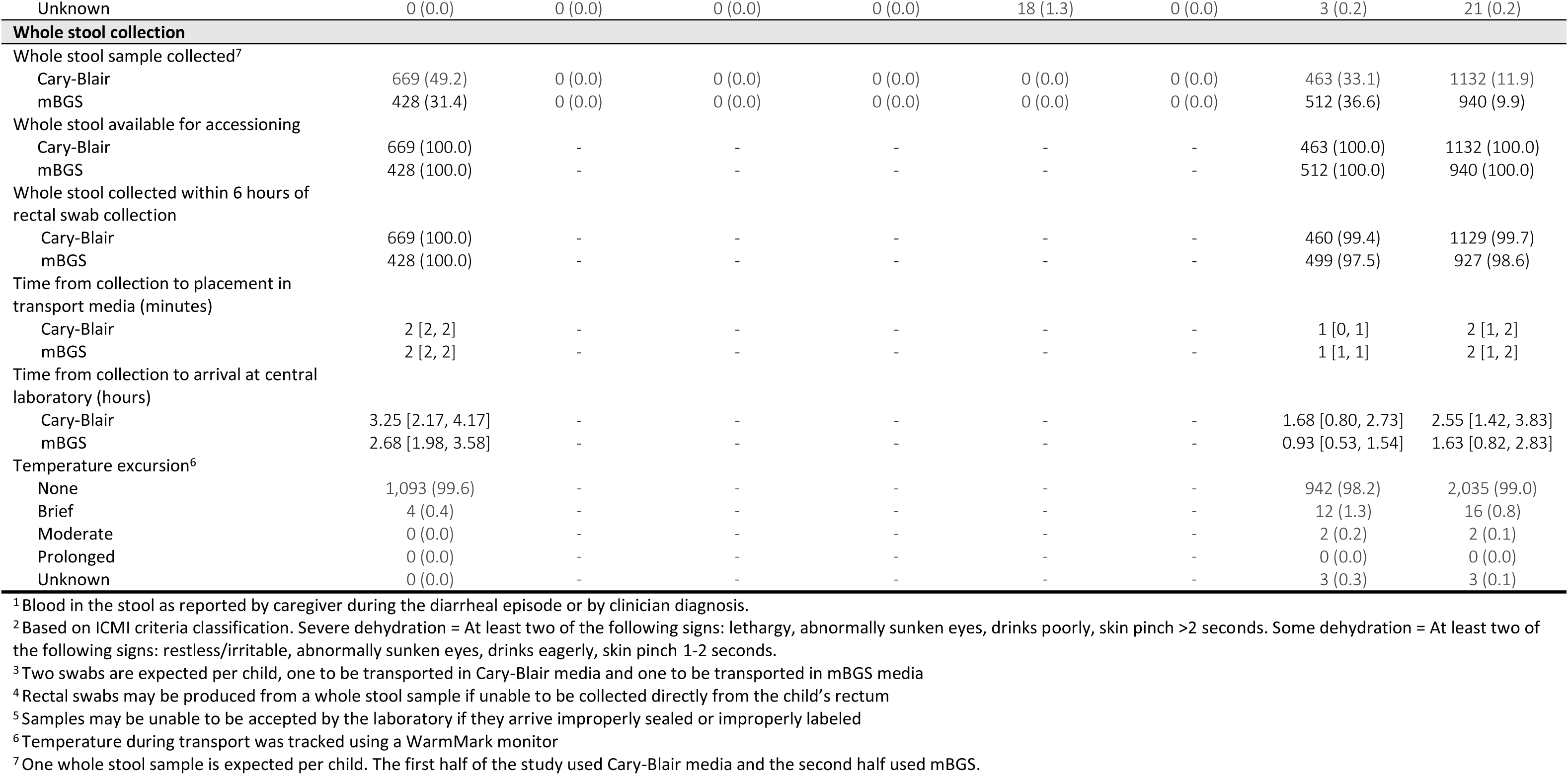
Participant characteristics and sample collection.

|  | Bangladesh | Kenya | Malawi | Mali | Pakistan | Peru | The Gambia | Overall |
| --- | --- | --- | --- | --- | --- | --- | --- | --- |
|  | n (%) or median [IQR] |  |  |  |  |  |  |  |
| Enrolled | 1361 | 1400 | 1399 | 1400 | 1400 | 1117 | 1399 | 9476 |
| **Participant demographics** |  |  |  |  |  |  |  |  |
| Age (months) |  |  |  |  |  |  |  |  |
| 6-11 | 572 (42.0) | 560 (40.0) | 478 (34.2) | 600 (42.9) | 454 (32.4) | 365 (32.7) | 446 (31.9) | 3,475 (36.7) |
| 12-23 | 588 (43.2) | 595 (42.5) | 627 (44.8) | 613 (43.8) | 646 (46.1) | 585 (52.4) | 718 (51.3) | 4,372 (46.1) |
| 24-35 | 201 (14.8) | 245 (17.5) | 294 (21.0) | 187 (13.4) | 300 (21.4) | 167 (15.0) | 235 (16.8) | 1,629 (17.2) |
| Female | 596 (43.8) | 637 (45.5) | 651 (46.5) | 643 (45.9) | 655 (46.8) | 488 (43.7) | 646 (46.2) | 4,316 (45.5) |
| **Clinical characteristics** |  |  |  |  |  |  |  |  |
| Dysentery<sup>1</sup> | 206 (15.1) | 148 (10.6) | 138 (9.9) | 93 (6.6) | 234 (16.7) | 170 (15.2) | 241 (17.2) | 1,230 (13.0) |
| Dehydration<sup>2</sup> |  |  |  |  |  |  |  |  |
| None | 1,191 (87.5) | 656 (46.9) | 1,348 (96.4) | 1,236 (88.3) | 1,185 (84.6) | 214 (19.2) | 1,310 (93.6) | 7,140 (75.3) |
| Some | 168 (12.3) | 685 (48.9) | 49 (3.5) | 154 (11.0) | 207 (14.8) | 897 (80.3) | 70 (5.0) | 2,230 (23.5) |
| Severe | 2 (0.1) | 59 (4.2) | 2 (0.1) | 10 (0.7) | 8 (0.6) | 6 (0.5) | 19 (1.4) | 106 (1.1) |
| Hospitalized | 125 (9.2) | 80 (5.7) | 15 (1.1) | 1 (0.1) | 25 (1.8) | 15 (1.3) | 102 (7.3) | 363 (3.8) |
| **Rectal swab collection** |  |  |  |  |  |  |  |  |
| Rectal swabs collected<sup>3</sup> |  |  |  |  |  |  |  |  |
| Cary-Blair | 1,361 (100.0) | 1,400 (100.0) | 1,399 (100.0) | 1,400 (100.0) | 1,400 (100.0) | 1,117 (100.0) | 1,399 (100.0) | 9,476 (100.0) |
| mBGS | 1,361 (100.0) | 1,400 (100.0) | 1,399 (100.0) | 1,400 (100.0) | 1,400 (100.0) | 1,117 (100.0) | 1,399 (100.0) | 9,476 (100.0) |
| Rectal swabs collected from whole stool sample instead<sup>4</sup> | 0 (0.0) | 0 (0.0) | 3 (0.2) | 2 (0.1) | 51 (3.6) | 2 (0.2) | 8 (0.6) | 66 (0.7) |
| Rectal swab available for accessions<sup>5</sup> |  |  |  |  |  |  |  |  |
| Cary-Bair | 1,361 (100.0) | 1,400 (100.0) | 1,399 (100.0) | 1,400 (100.0) | 1,400 (100.0) | 1,117 (100.0) | 1,399 (100.0) | 9,476 (100.0) |
| mBGS | 1,361 (100.0) | 1,400 (100.0) | 1,399 (100.0) | 1,400 (100.0) | 1,400 (100.0) | 1,117 (100.0) | 1,399 (100.0) | 9,476 (100.0) |
| Time from collection to placement in transport media (minutes) |  |  |  |  |  |  |  |  |
| Cary-Blair | 2 [1, 2] | 1 [1, 2] | 1 [1, 2] | 1 [0, 1] | 1 [1, 1] | 2 [1, 2] | 1 [0, 1] | 1 [1, 2] |
| mBGS | 1 [1, 2] | 1 [1, 2] | 1 [1, 1] | 0 [0, 1] | 1 [0, 1] | 2 [1, 2] | 1 [0, 1] | 1 [1, 1] |
| Time from collection to arrival at central laboratory (hours) |  |  |  |  |  |  |  |  |
| Cary-Blair | 3.25 [2.33, 4.17] | 3.88 [2.93, 4.82] | 4.35 [3.67, 4.97] | 4.08 [2.35, 5.63] | 2.32 [1.75, 3.05] | 2.58 [1.67, 3.37] | 1.43 [0.83, 2.43] | 3.13 [1.93, 4.30] |
| mBGS | 3.25 [2.33, 4.17] | 3.88 [2.93, 4.82] | 4.33 [3.67, 4.97] | 4.08 [2.36, 5.63] | 2.32 [1.75, 3.04] | 2.58 [1.67, 3.37] | 1.43 [0.83, 2.45] | 3.13 [1.95, 4.30] |
| Temperature excursion<sup>6</sup> |  |  |  |  |  |  |  |  |
| None | 1,355 (99.6) | 1,314 (93.9) | 1,396 (99.8) | 1,343 (95.9) | 1,268 (90.6) | 1,071 (95.9) | 1,372 (98.1) | 9,119 (96.2) |
| Brief | 6 (0.4) | 86 (6.1) | 3 (0.2) | 24 (1.7) | 103 (7.4) | 7 (0.6) | 20 (1.4) | 249 (2.6) |
| Moderate | 0 (0.0) | 0 (0.0) | 0 (0.0) | 33 (2.4) | 11 (0.8) | 39 (3.5) | 4 (0.3) | 87 (0.9) |
| Prolonged | 0 (0.0) | 0 (0.0) | 0 (0.0) | 0 (0.0) | 0 (0.0) | 0 (0.0) | 0 (0.0) | 0 (0.0) |
| Unknown | 0 (0.0) | 0 (0.0) | 0 (0.0) | 0 (0.0) | 18 (1.3) | 0 (0.0) | 3 (0.2) | 21 (0.2) |
| **Whole stool collection** |  |  |  |  |  |  |  |  |
| Whole stool sample collected<sup>7</sup> |  |  |  |  |  |  |  |  |
| Cary-Blair | 669 (49.2) | 0 (0.0) | 0 (0.0) | 0 (0.0) | 0 (0.0) | 0 (0.0) | 463 (33.1) | 1132 (11.9) |
| mBGS | 428 (31.4) | 0 (0.0) | 0 (0.0) | 0 (0.0) | 0 (0.0) | 0 (0.0) | 512 (36.6) | 940 (9.9) |
| Whole stool available for accessioning |  |  |  |  |  |  |  |  |
| Cary-Blair | 669 (100.0) | - | - | - | - | - | 463 (100.0) | 1132 (100.0) |
| mBGS | 428 (100.0) | - | - | - | - | - | 512 (100.0) | 940 (100.0) |
| Whole stool collected within 6 hours of rectal swab collection |  |  |  |  |  |  |  |  |
| Cary-Blair | 669 (100.0) | - | - | - | - | - | 460 (99.4) | 1129 (99.7) |
| mBGS | 428 (100.0) | - | - | - | - | - | 499 (97.5) | 927 (98.6) |
| Time from collection to placement in transport media (minutes) |  |  |  |  |  |  |  |  |
| Cary-Blair | 2 [2, 2] | - | - | - | - | - | 1 [0, 1] | 2 [1, 2] |
| mBGS | 2 [2, 2] | - | - | - | - | - | 1 [1, 1] | 2 [1, 2] |
| Time from collection to arrival at central laboratory (hours) |  |  |  |  |  |  |  |  |
| Cary-Blair | 3.25 [2.17, 4.17] | - | - | - | - | - | 1.68 [0.80, 2.73] | 2.55 [1.42, 3.83] |
| mBGS | 2.68 [1.98, 3.58] | - | - | - | - | - | 0.93 [0.53, 1.54] | 1.63 [0.82, 2.83] |
| Temperature excursion<sup>6</sup> |  |  |  |  |  |  |  |  |
| None | 1,093 (99.6) | - | - | - | - | - | 942 (98.2) | 2,035 (99.0) |
| Brief | 4 (0.4) | - | - | - | - | - | 12 (1.3) | 16 (0.8) |
| Moderate | 0 (0.0) | - | - | - | - | - | 2 (0.2) | 2 (0.1) |
| Prolonged | 0 (0.0) | - | - | - | - | - | 0 (0.0) | 0 (0.0) |
| Unknown | 0 (0.0) | - | - | - | - | - | 3 (0.3) | 3 (0.1) |

<sup>1</sup>Blood in the stool as reported by caregiver during the diarrheal episode or by clinician diagnosis.
<sup>2</sup>Based on ICMI criteria classification. Severe dehydration = At least two of the following signs: lethargy, abnormally sunken eyes, drinks poorly, skin pinch >2 seconds. Some dehydration = At least two of the following signs: restless/irritable, abnormally sunken eyes, drinks eagerly, skin pinch 1-2 seconds.
<sup>3</sup>Two swabs are expected per child, one to be transported in Cary-Blair media and one to be transported in mBGS media
<sup>4</sup>Rectal swabs may be produced from a whole stool sample if unable to be collected directly from the child's rectum
<sup>5</sup>Samples may be unable to be accepted by the laboratory if they arrive improperly sealed or improperly labeled
<sup>6</sup>Temperature during transport was tracked using a WarmMark monitor
<sup>7</sup>One whole stool sample is expected per child. The first half of the study used Cary-Blair media and the second half used mBGS.

There was no overall statistical evidence (p=0.5) of a difference in *Shigella* culture isolation proportion between CB (7.8%) and mBGS (7.9%) (**Table 2**) except in Bangladesh where the isolation proportion was higher in mBGS (12.9% vs. 14.3%, p = 0.001). There was also no major differences in *Shigella* isolation between the two transport media within *Shigella* serotypes except Bangladesh for *S*. *flexneri* (p=0.04) and *S*. *sonnei* (p=0.02).

**Table 2:** Comparison of *Shigella* culture positivity in Cary Blair and mBGS transport media stratified by site and serotype.

| *Shigella* culture positivity<sup>1</sup>: n (%) |  |  |  |  |  |  |
| --- | --- | --- | --- | --- | --- | --- |
| Country | N | Cary Blair |  | mBGS |  | p-value<sup>2</sup> |
|  |  | n | % (CI) | n | % (CI) |  |
| **All *Shigella* isolates** |  |  |  |  |  |  |
| Bangladesh | 1361 | 175 | 12.9 (11.2, 14.7) | 195 | 14.3 (12.6, 16.3) | 0.001 |
| Kenya | 1400 | 68 | 4.9 (3.8, 6.1) | 67 | 4.8 (3.8, 6.0) | >0.99 |
| Malawi | 1399 | 73 | 5.2 (4.2, 6.5) | 78 | 5.6 (4.5, 6.9) | 0.560 |
| Mali | 1400 | 69 | 4.9 (3.9, 6.2) | 65 | 4.6 (3.7, 5.9) | 0.571 |
| Pakistan | 1400 | 131 | 9.4 (7.9, 11.0) | 120 | 8.6 (7.2, 10.2) | 0.222 |
| Peru<sup>3</sup> | 843 | 64 | 7.6 (6.0, 9.6) | 65 | 7.7 (6.1, 9.7) | >0.99 |
| The Gambia | 1399 | 134 | 9.6 (8.1, 11.2) | 135 | 9.6 (8.2, 11.3) | >0.99 |
| **Overall** | **9202** | **714** | **7.8 (7.2, 8.3)** | **725** | **7.9 (7.3, 8.4)** | **0.545** |
| ***S. flexneri*** |  |  |  |  |  |  |
| Bangladesh | 1361 | 85 | 6.2 (5.1, 7.7) | 93 | 6.8 (5.6, 8.3) | 0.043 |
| Kenya | 1400 | 40 | 2.9 (2.1, 3.9) | 37 | 2.6 (1.9, 3.6) | 0.646 |
| Malawi | 1399 | 25 | 1.8 (1.2, 2.6) | 32 | 2.3 (1.6, 3.2) | 0.169 |
| Mali | 1400 | 47 | 3.4 (2.5, 4.4) | 44 | 3.1 (2.4, 4.2) | 0.606 |
| Pakistan | 1400 | 72 | 5.1 (4.1, 6.4) | 62 | 4.4 (3.5, 5.6) | 0.123 |
| Peru<sup>3</sup> | 843 | 45 | 5.3 (4.0, 7.1) | 46 | 5.5 (4.1, 7.2) | >0.99 |
| The Gambia | 1399 | 85 | 6.1 (4.9, 7.4) | 88 | 6.3 (5.1, 7.7) | 0.663 |
| **Overall** | **9202** | **399** | **4.3 (3.9, 4.8)** | **402** | **4.4 (4.0, 4.8)** | **0.872** |
| ***S. sonnei*** |  |  |  |  |  |  |
| Bangladesh | 1361 | 57 | 4.2 (3.2, 5.4) | 66 | 4.8 (3.8, 6.1) | 0.016 |
| Kenya | 1400 | 20 | 1.4 (0.9, 2.2) | 21 | 1.5 (1.0, 2.3) | >0.99 |
| Malawi | 1399 | 31 | 2.2 (1.6, 3.1) | 31 | 2.2 (1.6, 3.1) | >0.99 |
| Mali | 1400 | 11 | 0.8 (0.4, 1.4) | 11 | 0.8 (0.4, 1.4) | >0.99 |
| Pakistan | 1400 | 32 | 2.3 (1.6, 3.2) | 32 | 2.3 (1.6, 3.2) | >0.99 |
| Peru<sup>3</sup> | 843 | 19 | 2.2 (1.4, 3.5) | 19 | 2.2 (1.4, 3.5) | >0.99 |
| The Gambia | 1399 | 38 | 2.7 (2.0, 3.7) | 37 | 2.6 (1.9, 3.6) | >0.99 |
| **Overall** | **9202** | **208** | **2.3 (2.0, 2.6)** | **217** | **2.4 (2.1, 2.7)** | **0.356** |
| ***S. boydii*** |  |  |  |  |  |  |
| Bangladesh | 1361 | 24 | 1.8 (1.2, 2.6) | 23 | 1.7 (1.1, 2.5) | >0.99 |
| Kenya | 1400 | 5 | 0.4 (0.1, 0.8) | 6 | 0.4 (0.2, 0.9) | >0.99 |
| Malawi | 1399 | 10 | 0.7 (0.4, 1.3) | 7 | 0.5 (0.2, 1.0) | 0.450 |
| Mali | 1400 | 9 | 0.6 (0.3, 1.2) | 9 | 0.6 (0.3, 1.2) | >0.99 |
| Pakistan | 1400 | 15 | 1.1 (0.6, 1.8) | 15 | 1.1 (0.6, 1.8) | >0.99 |
| Peru<sup>3</sup> | 843 | 0 | 0 (-) | 0 | 0 (-) | - |
| The Gambia | 1399 | 8 | 0.6 (0.3, 1.1) | 7 | 0.5 (0.2, 1.0) | >0.99 |
| **Overall** | **9202** | **71** | **0.8 (0.6, 1.0)** | **67** | **0.7 (0.6, 0.9)** | **0.571** |
| ***S. dysenteriae*** |  |  |  |  |  |  |
| Bangladesh | 1361 | 7 | 0.5 (0.2, 1.1) | 10 | 0.7 (0.4, 1.4) | 0.248 |
| Kenya | 1400 | 0 | 0 (-) | 2 | 0.1 (0.0, 0.5) | - |
| Malawi | 1399 | 7 | 0.5 (0.2, 1.0) | 8 | 0.6 (0.3, 1.1) | >0.99 |
| Mali | 1400 | 2 | 0.1 (0.0, 0.5) | 1 | 0.1 (0.0, 0.4) | >0.99 |
| Pakistan | 1400 | 8 | 0.6 (0.3, 1.1) | 9 | 0.6 (0.3, 1.2) | >0.99 |
| Peru<sup>3</sup> | 843 | 0 | 0 (-) | 0 | 0 (-) | - |
| The Gambia | 1399 | 1 | 0.1 (0.0, 0.4) | 1 | 0.1 (0.0, 0.4) | - |
| **Overall** | **9202** | **25** | **0.3 (0.2, 0.4)** | **31** | **0.3 (0.2, 0.5)** | **0.149** |

CI: 95% confidence interval, mBGS: modified buffered glycerol saline.
<sup>1</sup> Culture *Shigella* culture positivity is defined as the percentage of enrolled participants with *Shigella* isolated from culture out of total participants with both rectal swabs cultured using both Cary-Blair and mBGS transport media.
<sup>2</sup> McNemar's test of superiority.
<sup>3</sup> Includes only participants enrolled after January 19, 2023 due to a laboratory error that made previous results incomparable across media types.

Despite there being no major differences in overall isolation proportions between the two media, the two swabs each transported in unique media did not always identify the same children as having *Shigella.* There were 131 (15.3%) cases of *Shigella* isolated from the CB swab and not the mBGS swab and 142 (16.6%) isolated from the mBGS swab and not the CB swab (out of the 856 *Shigella* culture confirmed cases identified in total, **Supplementary Table 1**). Irrespective of transport media, two swabs were superior at identifying children with *Shigella* compared with one: 747 (7.9%) of participants had *Shigella* isolated when considering only each child’s first swab, and 881 (9.3%) when both swabs were included (p<0.001, **Table 3)**.

**Table 3:** Country-specific and overall *Shigella* culture positivity from rectal swab and whole stool samples, across sites involved in the whole stool/rectal swab comparison sub-study matched by transport media.

| Country | N | Shigella culture positivity<sup>1</sup> |  |  |
| --- | --- | --- | --- | --- |
|  |  | Rectal swab: n (%)<sup>2</sup> | Whole stool: n (%)<sup>3</sup> | Difference: % (CI) |
| All Shigella isolates |  |  |  |  |
| Bangladesh | 1097 | 147 (13.40) | 156 (14.22) | -0.82 (-1.51, -0.13) |
| The Gambia | 951 | 107 (11.25) | 104 (10.94) | 0.32 (-0.52, 1.15) |
| Overall | 2048 | 254 (12.40) | 260 (12.70) | -0.29 (-0.83, 0.24) |
| S. flexneri |  |  |  |  |
| Bangladesh | 1097 | 70 (6.38) | 73 (6.65) | -0.27 (-0.72, 0.18) |
| The Gambia | 951 | 70 (7.36) | 66 (6.94) | 0.42 (-0.32, 1.16) |
| Overall | 2048 | 140 (6.84) | 139 (6.79) | 0.05 (-0.37, 0.47) |
| S. sonnei |  |  |  |  |
| Bangladesh | 1097 | 47 (4.28) | 53 (4.83) | -0.55 (-1.02, -0.07) |
| The Gambia | 951 | 29 (3.05) | 29 (3.05) | 0.00 (-0.25, 0.25) |
| Overall | 2048 | 76 (3.71) | 82 (4.00) | -0.29 (-0.57, -0.01) |
| S. boydii |  |  |  |  |
| Bangladesh | 1097 | 19 (1.73) | 18 (1.64) | 0.09 (-0.17, 0.35) |
| The Gambia | 951 | 5 (0.53) | 5 (0.53) | 0.00 (-0.25, 0.25) |
| Overall | 2048 | 24 (1.17) | 23 (1.12) | 0.05 (-0.13, 0.23) |
| S. dysenteriae |  |  |  |  |
| Bangladesh | 1097 | 9 (0.82) | 10 (0.91) | -0.09 (-0.24, 0.06) |
| The Gambia | 951 | 1 (0.11) | 2 (0.21) | -0.11 (-0.28, 0.07) |
| Overall | 2048 | 10 (0.49) | 12 (0.59) | -0.10 (-0.21, 0.02) |

CI: 90% confidence interval, mBGS: modified buffered glycerol saline, Prop: proportion.
<sup>1</sup> *Shigella* culture positivity is defined as the percentage of participants with *Shigella* isolated from culture out of total participants with both a rectal swab and a whole stool sample collected for the *Shigella* culture comparison.
<sup>2</sup> Includes only the first collected rectal swab from each child.
<sup>3</sup> Whole stool was collected among children who produce a sample while still at the enrollment facility.

Among the 2048 children with both whole stool and rectal swab samples, *Shigella* was isolated from 235 (11.5%) participants from both whole stool and rectal swab samples while 19 (0.09%) participants had *Shigella* isolated from rectal swab only and 25 (1.2%) from whole stool only (**Supplementary Table 2**). Rectal swabs were no worse than whole stool at isolating *Shigella* by culture (12.4% and 12.7%, respectively) with a difference of -0.29% (95% confidence interval -0.83% to 0.24 (**Table 4**). In site and serotype specific stratification, there was no evidence of non-inferiority in The Gambia but in Bangladesh we could not reject the null hypothesis that rectal swabs were non-inferior (13.40%) to whole stool (14.22%) (Difference: -0.82% [-1.51%, -0.13%]) among all species (**Table 4**). However, rectal swabs were non-inferior to whole stool in Bangladesh when both rectal swabs were compared to whole stool instead of only one swab (14.68% isolation vs. 14.22% isolation, respectively; difference 0.46% [95%CI: 0.06%, 0.85%], **Supplementary Table 3**) and remained non-inferior when subset by species. Including both of a child’s rectal swabs in the comparison to whole stool, 252 (12.3%) participants had *Shigella* isolated from both sample types, 26 (1.3%) were identified by rectal swab only, and 8 (0.04%) identified from whole stool only (**Supplementary Table 4**).

**Table 4:** Comparison of the timeline of stool processing procedures by *Shigella* culture positivity by site and *Shigella* spp.

|  | Bangladesh |  | Kenya |  | Malawi |  | Mali<br>Median (IQR) |  | Pakistan |  | Peru |  | The Gambia |  |
| --- | --- | --- | --- | --- | --- | --- | --- | --- | --- | --- | --- | --- | --- | --- |
| *Shigella* | - | + | - | + | - | + | - | + | - | + | - | + | - | + |
| **N** | **1166** | **195** | **1333** | **67** | **1321** | **78** | **1335** | **65** | **1280** | **120** | **1044** | **73** | **1264** | **135** |
| **Swab collection to placement in transport media (mins)** | 1.5<br>(1.5, 1.5) | 1.5<br>(1.5, 1.5) | 1.5<br>(1, 1.5) | 1.5<br>(1, 1.5) | 1<br>(0.5, 1.5) | 1<br>(0.625, 1.5) | 0.5<br>(0.5, 0.5) | 0.5<br>(0.5, 0.5) | 1<br>(0.5, 1.5) | 1<br>(0.5, 1.5) | 1.5<br>(1.5, 1.5) | 1.5<br>(1.5, 1.5) | 0.5<br>(0, 1) | 0.5<br>(0.5, 1) |
| **Transport media placement to placement in cool box (mins)** | 1.5<br>(1.5, 1.5) | 1.5<br>(1.5, 1.5) | 1.5<br>(1.5, 1.5) | 1.5<br>(1.5, 1.5) | 170.5<br>(140, 203.5) | 175.25<br>(143.5, 217.3) | 0.5<br>(0.5, 0.5) | 0.5<br>(0.5, 0.5) | 1.5<br>(1, 2.5) | 2<br>(1, 2.5) | 3.5<br>(2.5, 4.5) | 3.5<br>(2, 4) | 1<br>(0.5, 1.5) | 1<br>(0.5, 1.5) |
| **Placement in cool box to departure from clinic (mins)** | 137<br>(77, 187) | 133<br>(77, 194.5) | 106<br>(52, 172) | 97<br>(39, 163.5) | 5<br>(4, 10) | 6<br>(4, 10) | 161<br>(76, 255) | 178<br>(76, 254) | 85<br>(48, 128) | 84<br>(53.75, 117) | 112<br>(54.75, 160) | 122<br>(62, 153) | 68.5<br>(34, 128) | 68<br>(36, 115.5) |
| **Departure from clinic to arrival at central lab for inoculation (mins)** | 55<br>(35, 70) | 60<br>(40, 70) | 102<br>(67, 146) | 105<br>(68.5, 168) | 70<br>(54, 91) | 70<br>(50, 92.5) | 71<br>(46, 93) | 69<br>(45, 93) | 45<br>(30, 65) | 49<br>(33.75, 70) | 30<br>(20, 45) | 30<br>(15, 39) | 12<br>(9, 20) | 11<br>(7, 15) |

Abbreviations: IQR: interquartile range, mins: minutes

No meaningful differences were observed in *Shigella* isolation proportions from rectal swabs by timing between collection and placement in transport media, between collection and arrival at the central laboratory for primary plating, and by whether or not appropriate cold chain was maintained during specimen shipment (**Table 5**, **Figure 1 & 2**).

**Figure 1:**
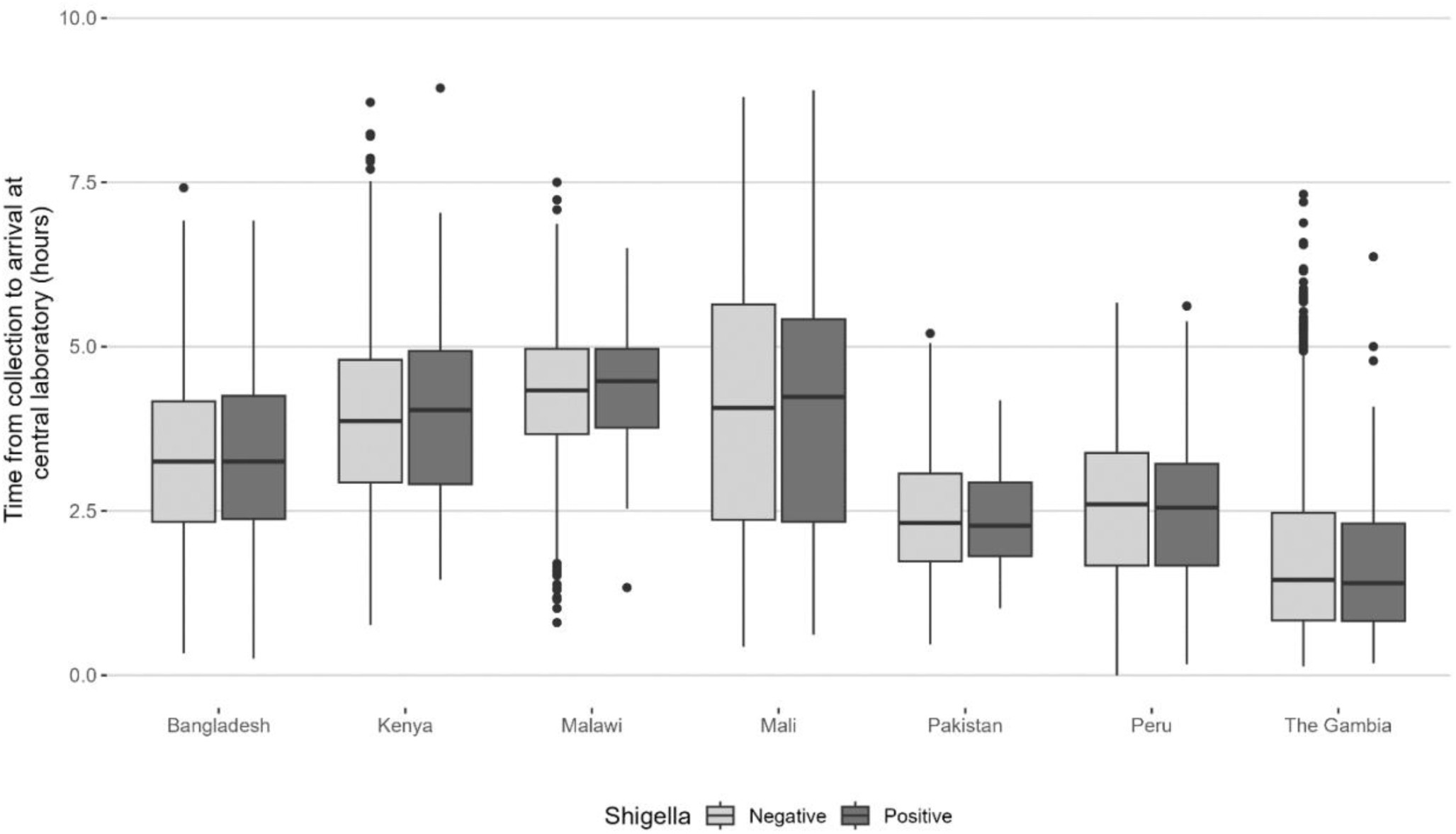
Comparison of time from sample collection to specimen arrival at the central laboratory and by *Shigella* positivity by culture.

**Figure 2:**
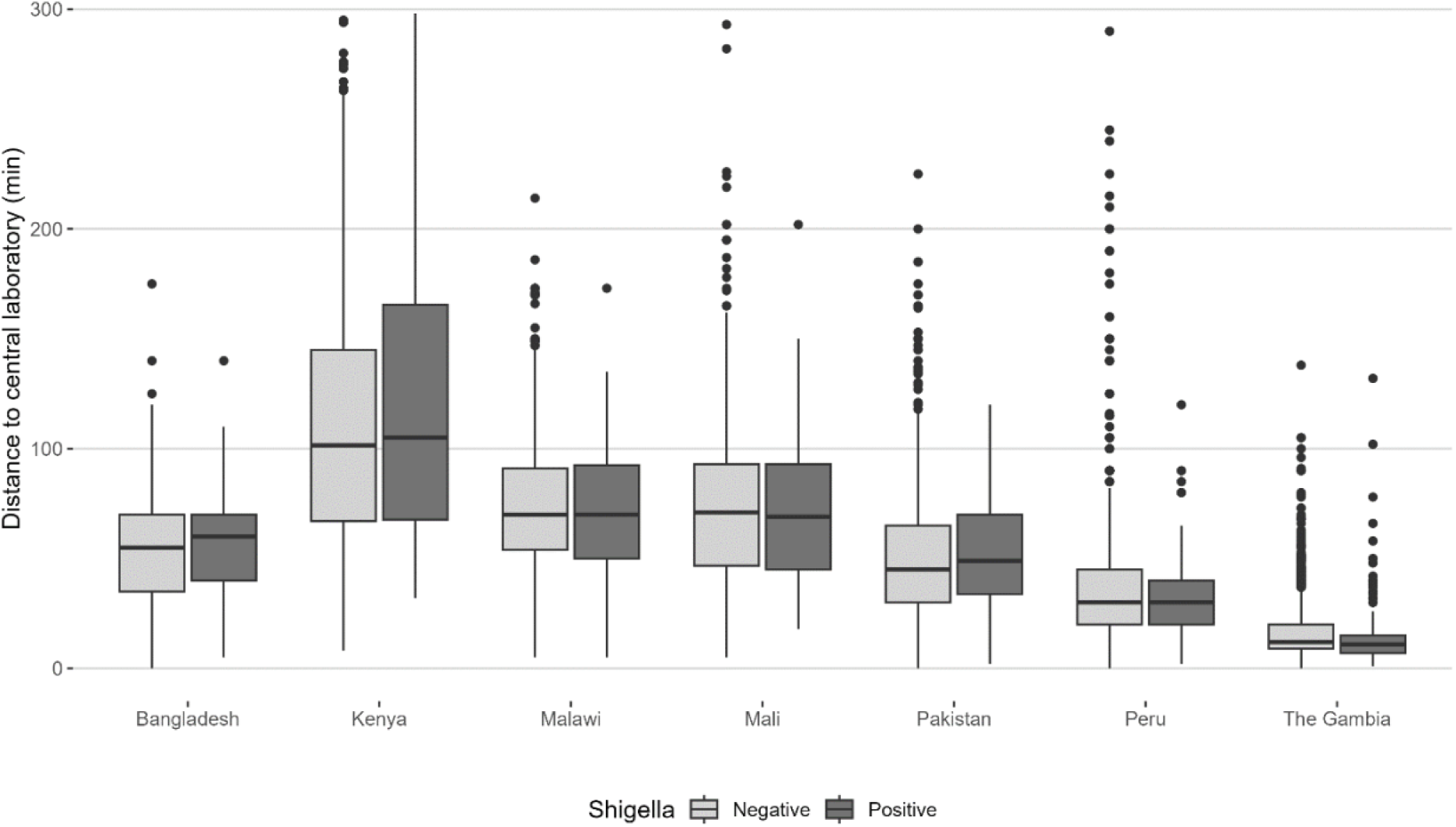
Comparison of *Shigella* culture positivity by distance from the clinic to central laboratory.

**Table 5:** Comparison of *Shigella* isolation comparing one swab sample to two swab samples.

|  |  | Shigella culture positivity<br>n (%) |  | Difference<br>% (CI) | p-value<sup>1</sup> |
| --- | --- | --- | --- | --- | --- |
|  | Enrolled | First swab | Either swab |  |  |
| All Shigella serotypes |  |  |  |  |  |
| Bangladesh | 1361 | 180 (13.2%) | 200 (14.7%) | -1.47 (-2.11, -0.83%) | <0.001 |
| Kenya | 1400 | 70 (5.0%) | 82 (5.9%) | -0.86 (-1.34, -0.37%) | 0.001 |
| Malawi | 1399 | 76 (5.4%) | 99 (7.1%) | -1.64 (-2.31, -0.98%) | <0.001 |
| Mali | 1400 | 70 (5.0%) | 81 (5.8%) | -0.79 (-1.25, -0.32%) | 0.001 |
| Pakistan | 1400 | 126 (9.0%) | 159 (11.4%) | -2.36 (-3.15, -1.56%) | <0.001 |
| Peru | 1117 | 93 (8.3%) | 114 (10.2%) | -1.88 (-2.68, -1.08%) | <0.001 |
| The Gambia | 1399 | 132 (9.4%) | 146 (10.4%) | -1.00 (-1.52, -0.48%) | <0.001 |
| Overall | 9476 | 747 (7.9%) | 881 (9.3%) | -1.41 (-1.65, -1.18%) | <0.001 |
| S. flexneri |  |  |  |  |  |
| Bangladesh | 1361 | 86 (6.3%) | 95 (7.0%) | -0.66 (-1.09, -0.23%) | 0.003 |
| Kenya | 1400 | 39 (2.8%) | 48 (3.4%) | -0.64 (-1.06, -0.22%) | 0.003 |
| Malawi | 1399 | 26 (1.9%) | 38 (2.7%) | -0.86 (-1.34, -0.37%) | 0.001 |
| Mali | 1400 | 47 (3.4%) | 53 (3.8%) | -0.43 (-0.77, -0.09%) | 0.014 |
| Pakistan | 1400 | 67 (4.8%) | 84 (6.0%) | -1.21 (-1.79, -0.64%) | <0.001 |
| Peru | 1117 | 65 (5.8%) | 80 (7.2%) | -1.34 (-2.02, -0.67%) | <0.001 |
| The Gambia | 1399 | 83 (5.9%) | 97 (6.9%) | -1.00 (-1.52, -0.48%) | <0.001 |
| Overall | 9476 | 413 (4.4%) | 495 (5.2%) | -0.87 (-1.05, -0.68%) | <0.001 |
| S. sonnei |  |  |  |  |  |
| Bangladesh | 1361 | 58 (4.3%) | 67 (4.9%) | -0.66 (-1.09, -0.23%) | 0.003 |
| Kenya | 1400 | 23 (1.6%) | 24 (1.7%) | -0.07 (-0.21, 0.07%) | 0.317 |
| Malawi | 1399 | 32 (2.3%) | 40 (2.9%) | -0.57 (-0.97, -0.18%) | 0.005 |
| Mali | 1400 | 11 (0.8%) | 13 (0.9%) | -0.14 (-0.34, 0.06%) | 0.157 |
| Pakistan | 1400 | 32 (2.3%) | 42 (3.0%) | -0.71 (-1.16, -0.27%) | 0.002 |
| Peru | 1117 | 28 (2.5%) | 33 (3.0%) | -0.45 (-0.84, -0.06%) | 0.025 |
| The Gambia | 1399 | 38 (2.7%) | 38 (2.7%) | 0.00 (0.00, 0.00%) | - |
| Overall | 9476 | 222 (2.3%) | 257 (2.7%) | -0.37 (-0.49, -0.25%) | <0.001 |
| S. boydii |  |  |  |  |  |
| Bangladesh | 1361 | 25 (1.8%) | 25 (1.8%) | 0.00 (0.00, 0.00%) | - |
| Kenya | 1400 | 6 (0.4%) | 6 (0.4%) | 0.00 (0.00, 0.00%) | - |
| Malawi | 1399 | 10 (0.7%) | 12 (0.9%) | -0.14 (-0.34, 0.06%) | 0.157 |
| Mali | 1400 | 10 (0.7%) | 13 (0.9%) | -0.21 (-0.46, 0.03%) | 0.083 |
| Pakistan | 1400 | 17 (1.2%) | 19 (1.4%) | -0.14 (-0.34, 0.06%) | 0.157 |
| Peru | 1117 | 0 (0.0%) | 0 (0.0%) | 0.00 (0.00, 0.00%) | - |
| The Gambia | 1399 | 8 (0.6%) | 8 (0.6%) | 0.00 (0.00, 0.00%) | - |
| Overall | 9476 | 76 (0.8%) | 83 (0.9%) | -0.07 (-0.13, -0.02%) | 0.008 |
| S. dysenteriae |  |  |  |  |  |
| Bangladesh | 1361 | 9 (0.7%) | 10 (0.7%) | -0.07 (-0.22, 0.07%) | 0.317 |
| Kenya | 1400 | 0 (0.0%) | 2 (0.1%) | -0.14 (-0.34, 0.06%) | - |
| Malawi | 1399 | 8 (0.6%) | 9 (0.6%) | -0.07 (-0.21, 0.07%) | 0.317 |
| Mali | 1400 | 2 (0.1%) | 2 (0.1%) | 0.00 (0.00, 0.00%) | - |
| Pakistan | 1400 | 8 (0.6%) | 10 (0.7%) | -0.14 (-0.34, 0.06%) | 0.157 |
| Peru | 1117 | 0 (0.0%) | 1 (0.1%) | -0.09 (-0.26, 0.09%) | - |
| The Gambia | 1399 | 1 (0.1%) | 1 (0.1%) | 0.00 (0.00, 0.00%) | - |
| Overall | 9476 | 28 (0.3%) | 35 (0.4%) | -0.07 (-0.13, -0.02%) | 0.008 |

<sup>1</sup>McNemar's test

## Discussion

*Shigella* is challenging to detect by microbiologic culture therefore optimizing conditions and sample types are critical to ensuring appropriate classification of *Shigella*-confirmed diarrhea. In this multi-country *Shigella* focused study, we found that two transport media, CB and mBGS, yield similar *Shigella* isolation proportions; that a single rectal swab is sufficient to isolate *Shigella* compared to whole stool and that two swabs yield optimal *Shigella* isolation proportions. We also found that there may be some site-specific nuances to optimal methods and likely unique practicalities that will be important considerations for eventual vaccine trials.

CB medium is the Center for Disease Control recommended transport media for rectal swabs, known for its optimal buffering capacity and nutrient composition which supports enteric pathogen preservation [25]. Pre-packaged flocked rectal swabs in CB medium have become commercially available which significantly improves reproducibility, scalability and reduces preparation time. However, these swabs are more expensive and can have a relatively short shelf life. We found CB was not superior to mBGS, and in fact, found mBGS to be superior to CB in isolating *S. flexneri* and *S. sonnei* in Bangladesh, which has also been shown previously [20]. Whereas CB mainly contains Sodium thioglycolate, Sodium chloride, mono and di-potassium phosphate, mBGS contains glycerol and saline which together help to maintain the stability of bacteria during transportation and provides better conservation of *Shigella* [20]. However, unlike CB, mBGS requires preparation and assembly which brings practical challenges. Although the individual reagents of mBGS media are available, rectal swabs in mBGS are not commercially available and must be prepared prior to use. While mBGS is cost-effective, these extra steps require additional coordination and standardization which may make it less practical for large-scale field studies.

*Shigella* culture positivity proportion from rectal swabs and whole stool samples collected across The Gambia and Bangladesh, were highly comparable. This has also been demonstrated by previous studies, [13, 15, 16], studies that have also highlighted the practical advantage of rectal swabs enabling quick sample collection rather than waiting for whole stool to be produced. Immediate sample collection is particularly advantageous for a bacterium such as *Shigella* which is likely to be impacted by antibiotic use which might be provided soon after presentation to a health facility [15]. At the Bangladesh site only, whole stool yielded slightly higher *Shigella* culture positivity, however this difference went away when two rectal swabs were used instead of one.

Isolation proportions are impacted by factors such as transport media type, shipment temperature, and time between collection and initial processing [20, 25-27]. These factors must be carefully balanced to preserve pathogen survival while considering cost-effectiveness and logistical challenges. Because EFGH followed strict protocols and QA/QC to ensure 2-8°C temperatures were maintained during shipment and that inoculation on the primary plate occurred less than 16 hours from sample collection, we had little variability in these circumstances to explore their impact on *Shigella* isolation. In addition, although standard operating procedures were followed across all sites, subtle differences in staff competency, processing time and environmental conditions could also have introduced bias. This study specifically focused on isolating *Shigella* to inform future vaccine trial protocols. However, future studies are necessary to evaluate the utility of these sample types and transport media for recovering multiple diarrheal bacterial pathogens.

The EFGH study demonstrated high culture isolation proportions of *Shigella* by thorough attention to proper sample collection, transit, and media. To maximize *Shigella* recovery by culture, while weighing practical considerations for eventual multi-country vaccine trials where standardization and minimizing missed *Shigella* cases will be critical, we recommend collection of 2 flocked rectal swabs, transported in CB or mBGS media, and strict adherence to optimal transit temperatures and times, all of which was feasible at EFGH sites.

## Author Contributions

MTB, JL, JJ, BH, AH, HB, VM, JO, LR, FQ, PPY, FNQ, SS, KK, ST, EH, JC, and OS conceptualized the manuscript and contributed to writing and editing. EF conducted the statistical analysis following the statistical analysis plan developed by the EFGH consortium. The EFGH protocol was co-developed by members of the EFGH consortium. All authors reviewed and approved the final manuscript.

## Enterics for Global Health Study (EFGH) Consortium Byline

Dilruba Ahmed, Raphael Anyango, Evans Apondi, Hannah E. Atlas, Alex O. Awuor, Mutsai Bakali, Bubacarr E. Ceesay, Bakary Conteh, Nigel A. Cunliffe, Irum Fatima, Marguerite Fenwood Hughes, Sean R. Galagan, Ensa Gitteh, Md Ismail Hossen, Sadia Islam, Samba Juma Jallow, Youssouf Keita, Mariama Keita, Margaret N. Kosek, Zubair Latif, Clement Lefu, Anya M. Lewin, Rebecca Maguire, Chimwemwe Mhango, Maureen Ndalama, Billy Ogwel, Uduma Uma Onwuchekwa, Tackeshy Pinedo Vasquez, Md Nazmul Hasan Rajib, S.M. Azadul Alam Raz, Francesca Schiaffino, Olivia Lang Schultes, Wagner Valentino Shapiama Lopez, Catherine Sonye, Kirkby D. Tickell, Lang Wang, Yingdi Wang, Loyda Fiorella Zegarra Paredes

## Acknowledgments

The authors thank the children who participated in these studies and their families, and the dedicated physicians, nurses, scientists, and staff at each study site for their dedication and outstanding performance of clinical and laboratory study activities.

## Data Sharing

The EFGH statistical analysis plan (https://clinicaltrials.gov/study/NCT06047821) and study protocol (https://academic.oup.com/ofid/issue/11/Supplement_1) were made publicly available. The datasets were deidentified and anonymized and will be publicly available upon publication of the manuscript.

## EFGH Laboratory Methods Manuscript – Tables & Figures

Analyst: Erika Feutz

*Includes all data collected for EFGH*

### PURPOSE

This document is intended to provide primary manuscript first and last authors with a comprehensive list of tables and figures that have been prepared by this paper’s assigned data analyst. This document is expected to accompany the manuscript with the manuscript referring to tables and figures presented in this document. Tables and figures here use the final EFGH data cut and can therefore be considered Tables and figures are formatted and ordered as they are expected to appear in the resulting manuscript.

### SUPPLEMENTARY MATERIALS

**Supplemental Table 1:**
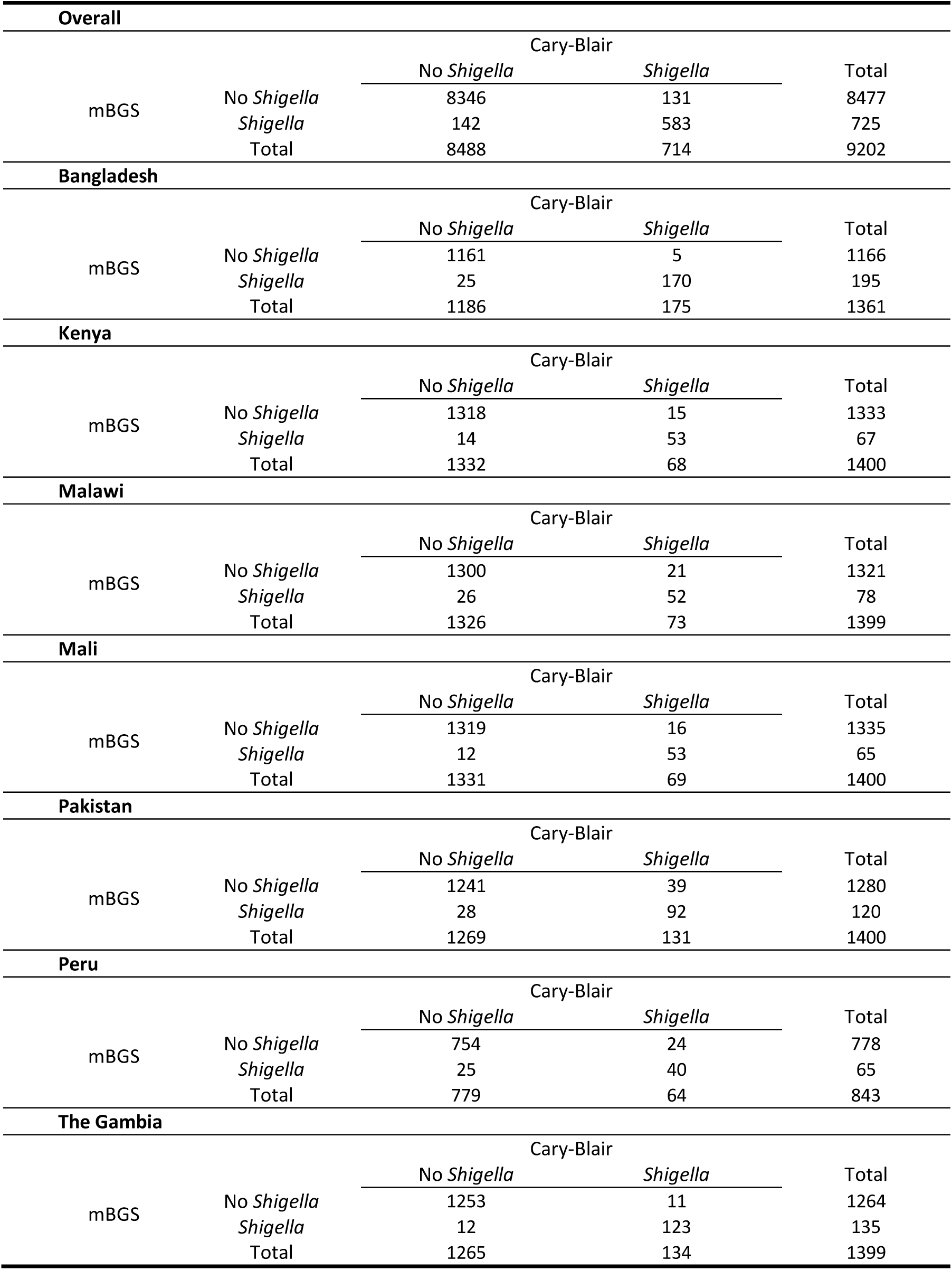
Concordance in *Shigella* culture positivity by media type (Cary-Blair vs. mBGS) Concordance in *Shigella* culture positivity by sample type (rectal swab vs. whole stool)

**Supplemental Table 2:**
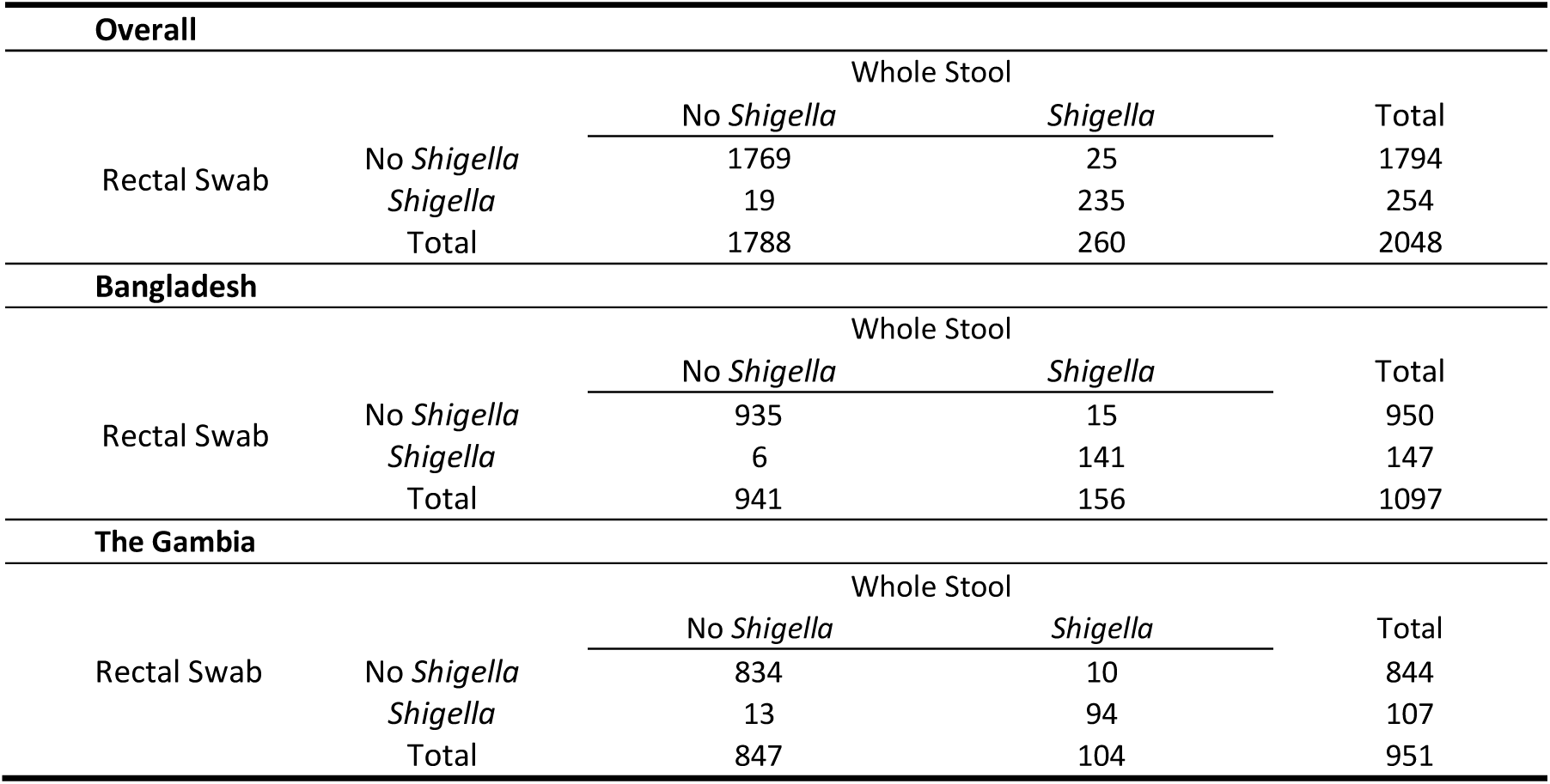
Two-by-two tables comparing *Shigella* positivity between rectal swab and whole stool using the first collected rectal swab matched with whole stool by media type.

**Supplemental Table 3:** Country-specific and overall *Shigella* culture positivity from rectal swab and whole stool samples, across sites involved in the whole stool/rectal swab comparison sub-study, including *Shigella* positivity in either rectal swab.

| Country | N | Shigella culture positivity<sup>1</sup> |  |  |
| --- | --- | --- | --- | --- |
|  |  | Rectal swab: n (%)<sup>2</sup> | Whole stool: n (%)<sup>3</sup> | Difference: % (CI) |
| All Shigella isolates |  |  |  |  |
| Bangladesh | 1097 | 161 (14.68) | 156 (14.22) | 0.46 (0.06, 0.85) |
| The Gambia | 951 | 117 (12.30) | 104 (10.94) | 1.37 (0.47, 2.27) |
| Overall | 2048 | 278 (13.57) | 260 (12.70) | 0.88 (0.41, 1.35) |
| S. flexneri |  |  |  |  |
| Bangladesh | 1097 | 76 (6.93) | 73 (6.65) | 0.27 (0.01, 0.53) |
| The Gambia | 951 | 80 (8.41) | 66 (6.94) | 1.47 (0.66, 2.28) |
| Overall | 2048 | 156 (7.62) | 139 (6.79) | 0.83 (0.43, 1.23) |
| S. sonnei |  |  |  |  |
| Bangladesh | 1097 | 54 (4.92) | 53 (4.83) | 0.09 (-0.17, 0.35) |
| The Gambia | 951 | 29 (3.05) | 29 (3.05) | 0.00 (-0.25, 0.25) |
| Overall | 2048 | 83 (4.05) | 82 (4.00) | 0.05 (-0.13, 0.23) |
| S. boydii |  |  |  |  |
| Bangladesh | 1097 | 19 (1.73) | 18 (1.64) | 0.09 (-0.17, 0.35) |
| The Gambia | 951 | 5 (0.53) | 5 (0.53) | 0.00 (-0.25, 0.25) |
| Overall | 2048 | 24 (1.17) | 23 (1.12) | 0.05 (-0.13, 0.23) |
| S. dysenteriae |  |  |  |  |
| Bangladesh | 1097 | 10 (0.91) | 10 (0.91) | 0.00 (0.00, 0.00) |
| The Gambia | 951 | 1 (0.11) | 2 (0.21) | -0.11 (-0.28, 0.07) |
| Overall | 2048 | 11 (0.54) | 12 (0.59) | -0.05 (-0.13, 0.03) |

CI: 90% confidence interval, mBGS: modified buffered glycerol saline, Prop: proportion.
<sup>1</sup> *Shigella* culture positivity is defined as the percentage of participants with *Shigella* isolated from culture out of total participants with both rectal swab and whole stool collected for the *Shigella* culture comparison.
<sup>2</sup> Includes *Shigella* positivity in either of two rectal swabs from each child.
<sup>3</sup> Whole stool was collected among children who produce a sample while still at the enrollment facility.

**Supplemental Table 4:**
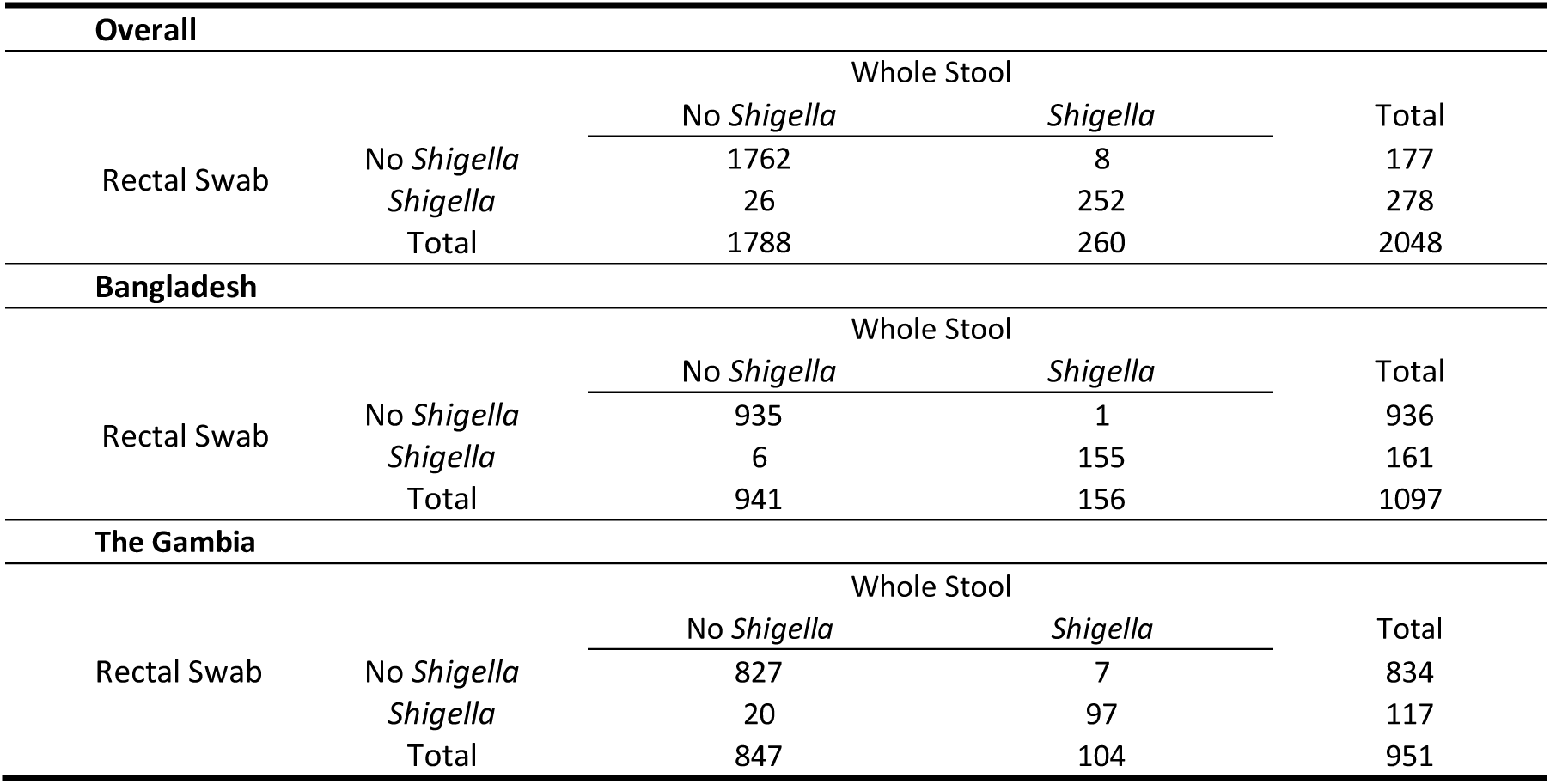
Two-by-two tables by site and *Shigella* species to compare *Shigella* positivity between rectal swab and whole stool using Shigella positivity of either rectal swab (matching the media type used for the same participant’s whole stool sample).

